# The Incidence of Immune Mediated Inflammatory Diseases Following COVID-19: a Matched Cohort Study in UK Primary Care

**DOI:** 10.1101/2022.10.06.22280775

**Authors:** Umer Syed, Anuradhaa Subramanian, David C Wraith, Janet M Lord, Kirsty McGee, Krishna Ghokale, Krishnarajah Nirantharakumar, Shamil Haroon

## Abstract

**Objective:** To assess whether there is an association between Severe Acute Respiratory Syndrome Coronavirus 2 (SARS CoV-2) infection and the incidence of immune mediated inflammatory diseases (IMIDs).

**Design:** Matched cohort study.

**Setting:** Primary care electronic health record data from the Clinical Practice Research Datalink Aurum database.

**Participants:** The exposed cohort included 458,147 adults aged 18 years and older with a confirmed SARS CoV-2 infection by reverse transcriptase polymerase chain reaction (RT-PCR) or lateral flow antigen test, and no prior diagnosis of IMIDs. They were matched on age, sex, and general practice to 1,818,929 adults in the unexposed cohort with no diagnosis of confirmed or suspected SARS CoV-2 infection and no prior diagnosis of IMIDs.

**Main Outcome Measures:** The primary outcome measure was a composite of the incidence of any of the following IMIDs: 1. autoimmune thyroiditis, 2. coeliac disease, 3. inflammatory bowel disease (IBD), 4. myasthenia gravis, 5. pernicious anaemia, 6. psoriasis, 7. rheumatoid arthritis (RA), 8. Sjogren’s syndrome, 9. systemic lupus erythematosus (SLE), 10. type 1 diabetes mellitus (T1DM), and 11. vitiligo. The secondary outcomes were the incidence of each of these conditions separately. Cox proportional hazards models were used to estimate adjusted hazard ratios (aHR) and 95% confidence intervals (CI) for the primary and secondary outcomes comparing the exposed to the unexposed cohorts, and adjusting for age, sex, ethnic group, smoking status, body mass index, relevant infections, and medications.

**Results:** 537 patients (0.11%) in the exposed cohort developed an IMID during the follow-up period over 0.29 person years, giving a crude incidence rate of 3.54 per 1000 person years. This was compared 1723 patients (0.09%) over 0.29 person years in the unexposed cohort, with an incidence rate of 2.82 per 1000 person years. Patients in the exposed cohort had a 22% relative increased risk of developing an IMID, compared to the unexposed cohort (aHR 1.22, 95% CI 1.10 to 1.34). The incidence of three IMIDs were statistically significantly associated with SARS CoV-2 infection. These were T1DM (aHR 1.56, 95% CI 1.09 to 2.23), IBD (1.52, 1.23 to 1.88), and psoriasis (1.23, 1.05 to 1.42).

**Conclusions:** SARS CoV-2 was associated with an increased incidence of IMIDs including T1DM, IBD and psoriasis. Further research is needed to replicate these findings in other populations and to measure autoantibody profiles in cohorts of individuals with COVID-19, including Long COVID and matched controls.

**Summary Box:** 

**What is already known on this topic:**

- A subsection of the population who tested positive for SARS CoV-2 is suffering from post-Covid-19 condition or long COVID.
- Preliminary findings, such as case reports of post-COVID-19 IMIDs, increased autoantibodies in COVID-19 patients, and molecular mimicry of the SARS-CoV-2 virus have given rise to the theory that long COVID may be due in part to a deranged immune response.

**What this study adds:**

- COVID-19 exposure was associated with a 22% relative increase in the risk of developing certain IMIDs, including type 1 diabetes mellitus, inflammatory bowel disease, and psoriasis.
- These findings provide further support to the hypothesis that a subgroup of Long Covid may be caused by immune mediated mechanisms.

## Introduction

Emerging in late 2019, Severe Acute Respiratory Syndrome Coronavirus-2 (SARS-CoV-2), the virus causing the Coronavirus Disease-2019 (COVID-19) pandemic, has of July 2022, resulted in over 6 million deaths worldwide.^1,2^ The acute presentation can range from being completely asymptomatic all the way to sepsis, organ failure and death.^3^ The effects of COVID-19 are not limited solely to acute infection but have also manifested in a series of post-acute sequelae commonly referred to as Long COVID or post COVID-19 condition.^4,5^ The World Health Organisation define this as symptoms occurring in people with a history of probable or confirmed SARS CoV-2 infection three months after the onset of COVID-19 that cannot be explained by an alternative diagnosis.^5,6^ With over a third of people with COVID-19 reporting persistent symptoms and over 1.7 million UK residents self-reporting to have the condition, Long COVID is emerging as a one of the major public health challenges of the modern era.^7,8^ Despite this, the pathogenesis behind the condition remains unclear.^9,10^

One potential theory regarding its pathogenesis is that SARS-CoV-2 infection causes an inappropriate immune response that leads to the varied symptoms of Long COVID. This theory arose from evidence of a marked and persistent increase of autoantibodies in patients with COVID-19 compared to their uninfected controls and possibly half of patients hospitalised with COVID-19 being transiently positive for anti-phospholipid (aPL) antibodies.^11,12^ Some of these autoantibodies were also deemed as potential risk factors for Long COVID.^13^ Multiple systematic reviews have also collated case reports of patients with a history of COVID-19 who have experienced deranged immune manifestations. Tang *et al*. found 187 reports and Novelli *et al*. found 382 reports of autoimmune-like phenomena following COVID-19.^14,15^ Among those with a history of COVID-19, one review reported thyroid dysfunction in up to 20% of patients, which is linked with B and T-cell autoimmunity.^16^ This reported autoimmunity may be due to the degree of homology existing between some human self-proteins and components of SARS-CoV-2, a phenomenon termed molecular mimicry.^17^ Molecular mimicry combined with the immune system dysregulation that occurs during SARS-CoV-2 infection may be the mechanism driving the development of immune mediated inflammatory diseases. Alternatively, the reaction could arise from tissue damage and the release of autoantigens as a result of SARS-CoV-2 infection.

This preliminary evidence has been derived largely from case-series, case-reports, small cohort studies or systematic reviews of these study types, which are weak study designs for ascertaining causal inference. Stronger study designs are needed that include appropriate control groups and large sample sizes. Furthermore, the data are drawn largely from patients with moderate or severe COVID-19, which underrepresents the mild or asymptomatic cases that make up most SARS CoV-2 infections and can also go on to develop Long COVID.^18^ To address these limitations, we conducted a retrospective matched cohort study using data from a large primary care database to assess the incidence of IMIDs in patients with SARS CoV-2 infection compared to matched individuals with no record of SARS CoV-2 infection.

## Methods

### Study Design and Data Source

A retrospective cohort study was undertaken using data extracted from the Clinical Practice Research Datalink Aurum database between 31^st^ of January 2020 and 30^th^ of June 2021. The CPRD Aurum database consists of routinely collected, pseudo-anonymised data from general practices across England.^19^ The data were extracted using the Data extraction for epidemiological research (DExtER) tool, which facilitates extraction based upon predefined parameters.^20^

### Study Population

Patients were eligible to enter the study if they were at least 18 years old at the study start date, had no prior history of IMIDs that were included in the primary outcome (see below), had an acceptable patient flag indicating provision of good quality data and if they were registered with an eligible general practice for at least 12 months to allow sufficient time for recording baseline information.

### Exposure

All patients with a SNOMED-CT coded diagnosis of either a positive reverse transcriptase polymerase chain reaction (RT-PCR) or lateral flow antigen test for SARS-CoV-2 were included in the exposed cohort, and the date of coded diagnosis was assigned as the index date. For each exposed patient, up to four patients were selected without a coded record of a positive RT-PCR or lateral flow antigen test, or a diagnosis of suspected or confirmed diagnosis of COVID-19, and matched on age, sex and registered general practice. This made up the unexposed cohort. The same index date of the exposed patients was assigned to the corresponding matched unexposed patients to avoid immortal time bias.^21^

### Outcomes

The primary outcome was a composite of the incidence of any of the following IMIDS: 1. autoimmune thyroiditis, 2. coeliac disease, 3. inflammatory bowel disease (IBD), 4. myasthenia gravis, 5. pernicious anaemia, 6. psoriasis, 7. rheumatoid arthritis (RA), 8. Sjogren’s syndrome, 9. systemic lupus erythematosus (SLE), 10. type 1 diabetes mellitus (T1DM), and 11. vitiligo. These conditions were selected as they cover a range of different systems and constitute many of the most prevalent IMIDs in the UK. Thus, the likelihood of observing an effect was not greatly diminished by the limited number of conditions. The secondary outcomes were the individual diseases included in the primary outcome, to discern which of these IMIDs, if any, had the strongest association with SARS CoV-2 infection. SNOMED-CT code lists used for the ascertainment of each IMID, as well as the exposure codes, are given at https://github.com/Umer-Syed/COVIDAutoimmune.

### Follow-up Period

Participants were followed-up from the index date to the end of follow-up. The end of follow-up was defined as the earliest of any of the following: a coded diagnosis of an IMID, date of death, study end date (30^th^ June 2021), date of practice de-registration, and date of the last practice contribution to the CPRD Aurum database.

## Covariates

Age, sex, BMI, smoking status, ethnicity, previous exposure to viral infections (Epstein Barr virus (EBV), human cytomegalovirus (CMV), human herpesvirus 6 (HHV-6), human T lymphotropic virus type 1 (HTLV-1), hepatitis C virus (HCV), influenza A virus, and parvovirus B19), and previous prescriptions of selected medications (procainamide, hydralazine, quinidine, and isoniazid) were considered as confounders. This is due to previous studies finding these variables to be associated with at least one of the outcome IMIDs and were thus adjusted for in the analysis.^22-34^

Age was divided into the following bands: 18 to 29, 30 to 39, 40 to 49, 50 to 59, 60 to 69, and ≥70 years. Ethnicity was identified through Snomed CT Codes and was classified into the following groups: white, South Asian, Black, mixed ethnicity and other. BMI was divided in accordance with the WHO categories: underweight (body mass index (BMI) <18.5kg/m^2^), normal weight (18.5-24.9kg/m^2^), overweight (25-29.9kg/m^2^) and obese (≥30kg/m^2^).^35^ Finally, smoking status categorised patients into those who currently smoke, ex-smokers and those who have never smoked. Where data was missing for ethnicity, smoking status and BMI, a separate ‘data missing’ category was used.

### Statistical Analysis

Baseline characteristics of patients stratified by their exposure status were summarised using mean and standard deviation or median and interquartile range for continuous variables and frequency and percentages for categorical variables. The number and percentage of each of the outcome events for the unexposed and exposed cohorts were reported and the crude incidence rates per 1000 person-years were calculated. Cox proportional hazards regression models were used to estimate the unadjusted and adjusted hazard ratios (HRs) with 95% confidence intervals (CI), for each of the outcomes among patients in the exposed and unexposed cohorts. P-values below 0.05 were considered statistically significant. The risk factor analysis also used the Cox model with regards to the development of the composite outcome but included only the exposed group. All analyses were conducted using Stata Version 17, the do-file for this is given at https://github.com/Umer-Syed/COVIDAutoimmune.

## Results

### Baseline Characteristics

We identified 458,147 patients with confirmed SARS CoV-2 infection and matched them to 1,818,929 without confirmed or suspected COVID-19. Table 1 displays the baseline characteristics of patients in both cohorts. Both groups had slightly more females than males (54.7% versus 45.3%, respectively). The mean age was 43.6 years (SD 17.1) in the exposed cohort and 42.8 (SD 18.0) in the unexposed cohort. A slightly larger proportion of the exposed cohort were of white and South Asian ethnicity compared to the unexposed group (64.4% versus 59.4%, and 12.2% versus 10.6%, respectively). However, the unexposed cohort had a slightly higher amount of missing ethnicity data (21.6% versus 16.2%, respectively). The mean BMI was similar between groups but there were slightly more current smokers in the unexposed cohort (26.5% versus 22.1%, respectively). Exposure to the selected infections and medications was similar between both groups.

**Table 1.** Baseline characteristics of the exposed and unexposed cohorts

| Characteristics | Exposed cohort<br>(n=458,147) | Unexposed cohort<br>(n=1,818,929) |
| --- | --- | --- |
| **Sex, n (%)** |  |  |
| Male | 207,514 (45.3) | 823,988 (45.3) |
| Female | 250,633 (54.7) | 994,941 (54.7) |
| **Age (years)** |  |  |
| Mean (SD) | 43.6 (17.1) | 42.8 (18.0) |
| **Age categories, n (%)** |  |  |
| 18-29 | 113,618 (24.8) | 478,811 (26.3) |
| 30-39 | 94,499 (20.6) | 377,117 (20.7) |
| 40-49 | 87,707 (19.1) | 348,335 (19.2) |
| 50-59 | 84,454 (18.4) | 327,230 (18.0) |
| 60-69 | 41,841 (9.1) | 153,227 (8.4) |
| ≥70 | 36,028 (7.9) | 134,209 (7.4) |
| **Ethnicity, n (%)** |  |  |
| White | 295,067 (64.4) | 1,079,656 (59.4) |
| South Asian | 55,792 (12.2) | 192,600 (10.6) |
| Black | 19,226 (4.2) | 86,040 (4.7) |
| Mixed Ethnicity | 7066 (1.5) | 34,134 (1.9) |
| Other | 6614 (1.4) | 33,873 (1.9) |
| Missing | 74,382 (16.2) | 392,626 (21.6) |
| **BMI** |  |  |
| Mean (SD) | 27.6 (6.3) | 26.8 (6.1) |
| Median (IQR) | 26.7 (23.3-30.9) | 25.8 (22.5-29.8) |
| **BMI Categories, n (%)** |  |  |
| Underweight (<18.5 kg/m<sup>2</sup>) | 12,038 (2.6) | 59,644 (3.3) |
| Normal weight (18.5-25 kg/m<sup>2</sup>) | 136,480 (29.8) | 594,657 (32.7) |
| Overweight (25-30 kg/m<sup>2</sup>) | 131,076 (28.6) | 473,031 (26.0) |
| Obese (>30 kg/m<sup>2</sup>) | 116,836 (25.5) | 366,198 (20.1) |
| Missing | 61,717 (13.5) | 325,379 (17.9) |
| **Smoking Status, n (%)** |  |  |
| Never smoked | 168,415 (36.8) | 657,775 (36.2) |
| Ex-smoker | 167,507 (36.6) | 548,695 (30.2) |
| Current smoker | 101,351 (22.1) | 482,322 (26.5) |
| Missing | 20,874 (4.6) | 130,137 (7.2) |
| **Exposure to selected infection, n (%)** |  |  |
| All infections | 2354 (0.5) | 9098 (0.5) |
| EBV | 153 (0.0) | 579 (0.0) |
| CMV | 123 (0.0) | 327 (0.0) |
| HHV-6 | 0 (0.0) | 4 (0.0) |
| HTLV-1 | 0 (0.0) | 10 (0.0) |
| Hepatitis C virus | 784 (0.2) | 4 161 (0.2) |
| Influenza A virus | 968 (0.2) | 2 911 (0.2) |
| Parvovirus B19 | 282 (0.1) | 947 (0.1) |
| **Exposure to selected medication, n (%)** |  |  |
| All medications | 821 (0.2) | 2 552 (0.1) |
| **Procainamide** | 0 (0.00) | 2 (0.0) |
| **Hydralazine** | 218 (0.1) | 753 (0.0) |
| **Quinidine** | 26 (0.0) | 92 (0.0) |
| **Isoniazid** | 581 (0.1) | 1724 (0.1) |

### Primary Analysis

There were 537 (0.11%) outcomes observed among patients within the exposed cohort compared to 1723 (0.09%) within the unexposed cohort. The median (IQR) follow up was 0.29 years (0.24-0.42) for both groups. The results for the primary analysis are reported in Table 2 and depicted within Figure 1. The crude incidence rate (IR) per 1000 person-years was higher for the exposed cohort than the unexposed cohort (3.54 versus 2.82 per 1000 person-years, respectively). This yielded a crude hazard ratio of 1.26 (95% CI 1.14-1.39) for the composite primary outcome. When adjusted for covariates, the HR slightly reduced to 1.22 (1.10-1.34) but remained statistically significant. The median (IQR) follow up was 0.29 years (0.24-0.42) for both groups. The results for the primary analysis are detailed in Table 2 and shown visually within Figure 1.

**Table 2.** Incidence rates and HRs for the composite outcome

| Outcome | Exposed cohort<br>(n=458,147) | Unexposed cohort<br>(n=1,818,929) |
| --- | --- | --- |
| **Outcome events, No. (%)** | 537 (0.11) | 1 723 (0.09) |
| **Person-years of follow up** | 151568 | 611211 |
| **Crude incidence rate per 1000 person-years** | 3.54 | 2.82 |
| **Median follow-up, years (IQR)** | 0.29 (0.24-0.42) | 0.29 (0.24-0.42) |
| **Crude HR (95% CI)** | 1.26 (1.14-1.39) |  |
| **Adjusted\* HR (95% CI)** | 1.22 (1.10-1.34) |  |

\* adjusted for age, sex, BMI, ethnicity, smoking status, selected viral infections, and selected medications.
CI=confidence interval, HR=hazard ratio, IQR=interquartile range

**Figure 1:**
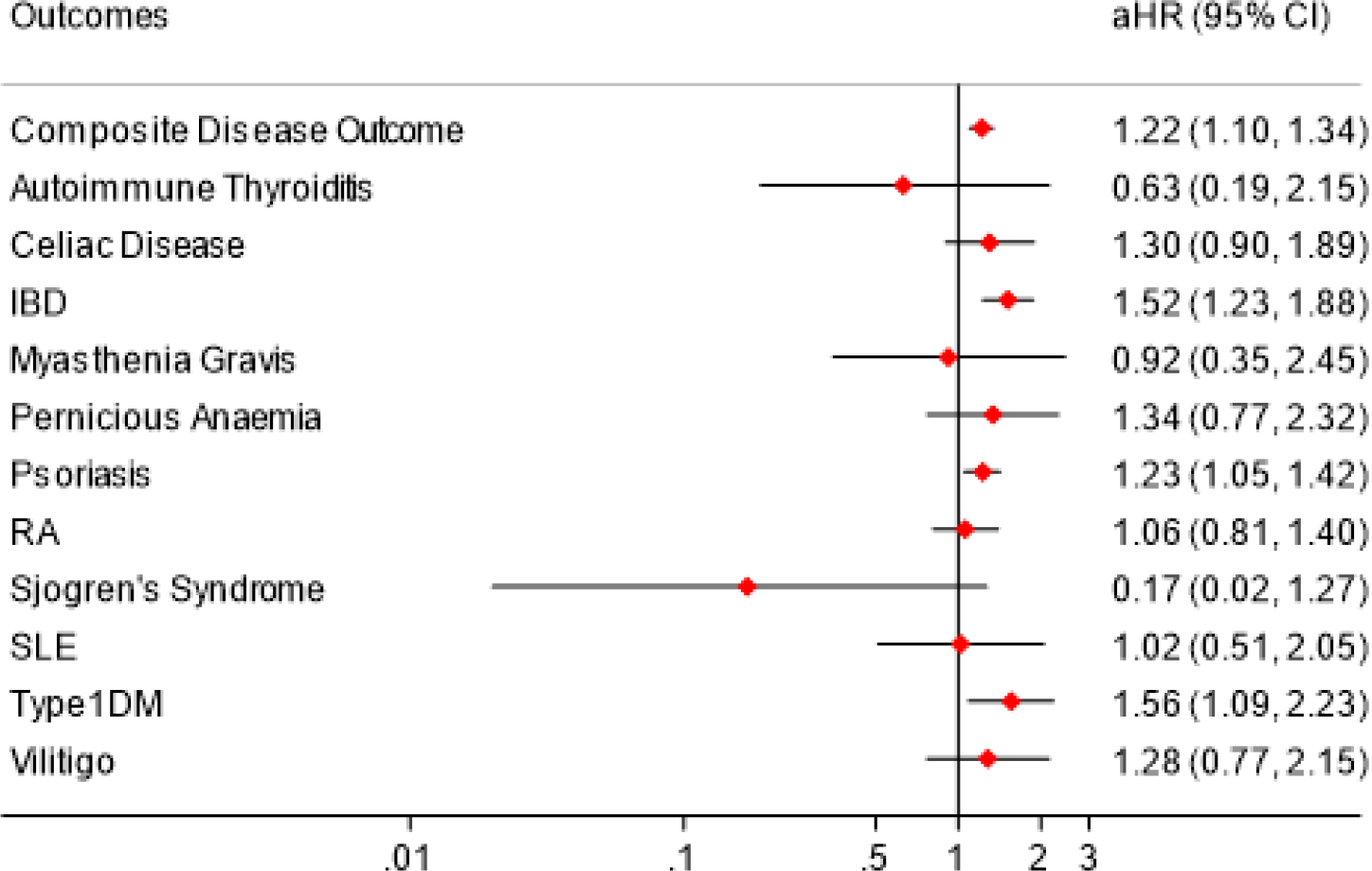
Forest plot of Adjusted HRs for IMIDs *aHR = Adjusted Hazard Ratio, CI = Confidence Interval, IBD = inflammatory bowel disease, RA = rheumatoid arthritis, SLE = systemic lupus erythematous, Type1DM = type 1 diabetes mellitus

### Secondary Analysis

Table 3 and Figure 2 report the results of each individual IMID as a separate outcome. Of the eleven conditions, SARS CoV-2 infection was significantly associated with an increased incidence of T1DM, IBD and psoriasis. T1DM was 56% more likely to occur in the exposed cohort compared the unexposed cohort (aHR 1.56, 95% CI 1.09 to 2.23). IBD was 52% more likely to occur in the exposed cohort compared to the unexposed cohort in relative terms (aHR 1.52, 95% CI 1.23 to 1.88). This was the second most common IMID to be diagnosed during the study period (21.6% of all IMIDs diagnosed in the exposed cohort and 17.9% in the unexposed cohort). Psoriasis was 23% more likely to occur in the exposed cohort compared to the unexposed cohort (1.23, 1.05 to 1.42) and was the most diagnosed IMID, representing more than 40% of all new diagnoses of IMIDs in both cohorts.

**Table 3.** Incidence rates and hazard ratios for individual IMIDs.

| Outcome | Exposed cohort | Unexposed cohort |
| --- | --- | --- |
| **Autoimmune thyroiditis** |  |  |
| Outcome events, No. (%*) | 3 (0.6) | 18 (1.0) |
| Crude incidence rate per 1000 person-years | 0.020 | 0.029 |
| Crude HR (95% CI) | 0.67(0.20-2.27) |  |
| Adjusted HR (95% CI) | 0.63 (0.19-2.15) |  |
| **Coeliac disease** |  |  |
| Outcome events, No. (%*) | 37 (6.9) | 113 (6.7) |
| Crude incidence rate per 1000 person-years | 0.24 | 0.18 |
| Crude HR (95% CI) | 1.32 (0.91-1.92) |  |
| Adjusted HR (95% CI) | 1.30 (0.90-1.89) |  |
| **Inflammatory bowel disease** |  |  |
| Outcome events, No. (%*) | 116 (21.6) | 309 (17.9) |
| Crude incidence rate per 1000 person-years | 0.77 | 0.51 |
| Crude HR (95% CI) | 1.51 (1.22-1.87) |  |
| Adjusted HR (95% CI) | 1.52 (1.23-1.88) |  |
| **Myasthenia gravis** |  |  |
| Outcome events, No. (%*) | 5 (0.9) | 21 (1.2) |
| Crude incidence rate per 1000 person-years | 0.033 | 0.034 |
| Crude HR (95% CI) | 0.96 (0.36-2.55) |  |
| Adjusted HR (95% CI) | 0.92 (0.35-2.45) |  |
| **Pernicious anaemia** |  |  |
| Outcome events, No. (%*) | 17 (3.2) | 50 (2.9) |
| Crude incidence rate per 1000 person-years | 0.11 | 0.082 |
| Crude HR (95% CI) | 1.37 (0.79-2.38) |  |
| Adjusted HR (95% CI) | 1.34 (0.77-2.32) |  |
| **Psoriasis** |  |  |
| Outcome events, No. (%*) | 223 (41.5) | 743 (43.1) |
| Crude incidence rate per 1000 person-years | 1.47 | 1.22 |
| Crude HR (95% CI) | 1.21 (1.04-1.41) |  |
| Adjusted HR (95% CI) | 1.23 (1.05-1.42) |  |
| **Rheumatoid arthritis** |  |  |
| Outcome events, No. (%*) | 66 (12.3) | 248 (14.4) |
| Crude incidence rate per 1000 person-years | 0.44 | 0.41 |
| Crude HR (95% CI) | 1.08 (0.82-1.41) |  |
| Adjusted HR (95% CI) | 1.06 (0.81-1.40) |  |
| **Sjogren's syndrome** |  |  |
| Outcome events, No. (%*) | 1 (0.2) | 23 (1.3) |
| Crude incidence rate per 1000 person-years | 0.00066 | 0.038 |
| Crude HR (95% CI) | 0.18 (0.02-1.31) |  |
| Adjusted HR (95% CI) | 0.17 (0.02-1.27) |  |
| **Systemic lupus erythematosus** |  |  |
| Outcome events, No. (%*) | 10 (1.9) | 39 (2.7) |
| Crude incidence rate per 1000 person-years | 0.066 | 0.064 |
| Crude HR (95% CI) | 1.03 (0.51-2.06) |  |
| Adjusted HR (95% CI) | 1.02 (0.51-2.05) |  |
| **Type 1 diabetes mellitus** |  |  |
| Outcome events, No. (%*) | 42 (7.8) | 106 (6.2) |
| Crude incidence rate per 1000 person-years | 0.28 | 0.17 |
| Crude HR (95% CI) | 1.60 (1.12-2.29) |  |
| Adjusted HR (95% CI) | 1.56 (1.09-2.23) |  |
| **Vitiligo** |  |  |
| Outcome events, No. (%*) | 19 (3.5) | 60 (3.5) |
| Crude incidence rate per 1000 person-years | 0.13 | 0.098 |
| Crude HR (95% CI) | 1.28 (0.76-2.14) |  |
| Adjusted HR (95% CI) | 1.28 (0.77-2.15) |  |

\* of total outcome events
CI=confidence interval, HR=hazard ratio

### Risk Factor Analysis

The results for the risk factor analysis are contained within Supplementary Table 1. A significant association was found with regards to sex and age. Variations in smoking status, ethnicity, BMI and exposure to selected infection or medication did not significantly impact the HR.

Females were 30% more likely to be diagnosed with an IMID than males (aHR 1.30, 95% CI 1.09 to 1.56). The risk also varied by age, with 40–49-year-olds and 50–59-year-olds being less likely to be diagnosed with an IMID than 18–29-year-olds (0.75, 0.58 to 0.99, and 0.69, 0.52 to 0.91, respectively). No other significant associations were found for the other risk factors considered in the model. No result was reported with regards to selected medication exposure. This is due to the confidence intervals being too large as there were a very small number of patients prescribed with the relevant medications. This resulted in an error where the STATA software was not able to calculate the 95% CI.

## Discussion

### Main Findings

Exposure to SARS CoV-2 infection was associated with a 22% relative increase in the incidence of any of the eleven IMIDs considered in our study compared to matched controls during the same period. This was after adjustment for several important confounding factors and during a relatively short period of follow up. We also found that this association was specific to an increased incidence of T1DM, inflammatory bowel disease, and psoriasis in the SARS CoV-2 infected cohort compared to the unexposed cohort.

### Comparison with Existing Literature

The relatively high incidence of psoriasis in the SARS CoV-2 infected cohort is supported by other reports from the literature which found that increased cases of psoriasis, and flares of existing disease, have been reported following COVID-19^.14^ Evidence regarding the development of IBD following COVID-19 is scarcer, although ulcerative colitis has been reported to develop post infection.^14^ A systematic review regarding T1DM and COVID-19 noted that between 1.77% and 15.6% of newly diagnosed patients, depending on the study, had preceding COVID-19^.36^

Female sex was also an important risk factor for incidence of IMIDs in our study. This is in line with previous evidence that female sex is a risk factor for most IMIDs and interestingly aligns with the consistent observation that women are at a higher risk of developing Long COVID than men^.37-39^ All age groups older than 30 years had a relatively lower incidence of IMIDs than the 18- to 29-year-old group, although this was only statistically significant in the 40- to 49-year-old and 50- to 59-year-old groups. The relationship between decreasing age and an increased risk of Long COVID has been found in another study, however, some studies do find an association between increasing age and Long COVID.^39-41^ This relationship may be due to the earlier age of diagnosis of T1DM, IBD and psoriasis, which made up the majority of outcome events in this study.^42-44^

Both smoking status and BMI were not significantly associated with the incidence of IMIDs, which conflicts with their presence as a known risk factor for both IMIDs in general and for Long COVID^.28,37,40^ However, the risk for developing IMIDs was significantly higher for overweight and obese patients with an increase of 4% and 18% respectively, compared to those with normal weight. A significant increase in risk was also found for current and ex-smokers with a 21% and 18% respective increase when compared to never smokers. The lack of statistically significant associations may have been due to an insufficient sample size or the short follow up period to adequately assess these risk factors.

### Strengths and Limitations

Data from over two million patients were included, which provided sufficient statistical power to assess for differences in the incidence of IMIDs between the exposed and unexposed cohorts over a relatively short follow-up period. This also allowed us to assess the relative incidence of eleven of the more common IMIDs across the two comparison groups. We included IMIDs in our outcome such as T1DM, that are likely to be well recorded in primary care records. The use of primary care data meant that we were able to adjust for important demographic and clinical risk factors that are known to be associated with the incidence of IMIDs. Use of data from practices across a national database also improved the generalisability of our findings. The use of primary care data also meant that numerous demographic factors, such as age, sex and ethnicity, were recorded. This facilitated covariate adjustment and risk factor analysis.

The study had several limitations. We had missing data for ethnicity (21% missing), BMI (17%), and smoking status (7%), which we accounted for in our analyses using a missing category variable. We did not have access to data on socioeconomic status but partially accounted for this by matching patients in the unexposed and exposed cohorts on general practice, which would result in patients from both groups sharing their residential geography and social demography. There is likely to be a degree of misclassification bias between the exposed and unexposed cohorts. There was little community testing for SARS CoV-2 infection in the first wave of the pandemic, which leaves open the possibility that some members of the unexposed cohort may have been infected but not diagnosed.

It is also likely that only a portion of the true number of diagnoses of IMIDs were detected during the study period with perhaps only the more severe cases being diagnosed. This is due to the relative inaccessibility of health services during the pandemic and the short period of follow-up in our study. The study period encompassed three national lockdowns where healthcare appointments were reduced leading to a backlog of up to 300,000 patients waiting over a year for treatment^.45,46^ The short follow up period may have diluted the effect size and power of the study as IMIDs typically can take a prolonged period of time to be diagnosed after the beginning of immune dysregulation and thus the full scope of the potential impact of SARS CoV-2 infection is likely to have been underrepresented^.47^ However, there is also the possibility that patients experiencing COVID-19 may have accessed healthcare services more than those with no prior infection, and thus had more opportunities to be diagnosed with IMIDs.

### Implications for Practice, Policy, and Research

Our findings provide epidemiological evidence that SARS CoV-2 infection is associated with an increased risk of a range of IMIDs, including T1DM, IBD, and psoriasis. This provides evidence that autoimmunity may be a potential mechanism that accounts for some of the longer-term symptoms and health impacts of a subgroup of those with Long COVID. This is particularly of interest given the finding that women may be at increased risk of both IMIDs as well as Long COVID, that symptoms of Long COVID are diverse and often overlap with those of IMIDs, and that the symptoms of both IMIDs and Long COVID characteristically follow a relapsing remitting pattern over time.^41^

However, there is currently a scarcity of studies assessing the relationship between SARS CoV-2 infection and the incidence of IMIDs. Further epidemiological studies with a longer follow-up period are needed to confirm our findings, and to test for relevant autoantibodies in the serum of participants to correlate with symptoms and clinical findings. These studies could also include other rarer IMIDs potentially associated with COVID-19 such as Guillain-Barre syndrome.^15^ Evidence suggests that those who have been vaccinated against COVID-19 are approximately half as likely to develop symptoms lasting over 28 days than unvaccinated individuals^.48^ It would be valuable to know if these differences in Long COVID incidence rates are also associated with differences in the incidence of IMIDs.

## Conclusion

SARS CoV-2 infection was associated with an increased incidence of several IMIDs, including type 1 diabetes mellitus, inflammatory bowel disease, and psoriasis. This finding lends support to the hypothesis that the long-term effects of COVID-19 or Long COVID may in part be related to increased autoimmunity. Further research is needed to replicate these findings in other populations and to sample autoantibody profiles in people with Long COVID as well as matched controls.

## Supporting information

RECORD checklist

## Data Availability

Data availability statement: Access to anonymized patient data from CPRD is subject to a data sharing agreement containing detailed terms and conditions of use following protocol approval from the MHRA Independent Scientific Advisory Committee. Details about Independent Scientific Advisory Committee applications and data costs are available on the CPRD website (cprd.com).

## Notes

### Competing Interest Statement

SH reports receiving funding from NIHR and UKRI. US and AS report no competing interests. JL receives grant funding from NIHR, UKRI, Versus Arthritis, The Scar Free Foundation, FOREUM, Bayer Healthcare for which also she acts as a consultant.

### Funding Statement

This research received no specific grant from any funding agency in the public, commercial, or not-for-profit sectors.

### Author Declarations

The project was reviewed and approved by the Clinical Practice Research Datalink (CPRD) Independent Scientific Advisory Committee (study #21_000712) on the 24th February 2022.

