## Supplementary Tables for "The Incidence of Immune Mediated Inflammatory Diseases Following COVID-19: a Matched Cohort Study in UK Primary Care"

**Supplementary Table 1** Adjusted hazard ratios for other risk factors included in the Cox proportional hazards model

| **Characteristics** | **Adjusted Hazard Ratio (95%CI)** |
| --- | --- |
| **SARS CoV-2 infection** | 1.22 (1.10-1.34) |
| **Sex** |  |
| **Male** | 1.00 (reference) |
| **Female** | 1.30 (1.09-1.56) |
| **Age** |  |
| **18-29 years old** | 1.00 (reference) |
| **30-39 years old** | 0.82 (0.64-1.06) |
| **40-49 years old** | 0.75 (0.58-0.99) |
| **50-59 years old** | 0.69 (0.52-0.91) |
| **60-69 years old** | 0.71 (0.50-1.01) |
| **≥70 years old** | 0.73 (0.50-1.05) |
| **Ethnicity** |  |
| **White** | 1.00 (reference) |
| **South Asian** | 0.97 (0.74-1.27) |
| **Black** | 0.65 (0.38-1.11) |
| **Mixed Ethnicity** | 0.35 (0.11-1.09) |
| **Other** | 0.94 (0.44–1.98) |
| **Missing** | 1.08 (0.86-1.37) |
| **Body mass index** |  |
| **Underweight (<18.5 kg/m^2^)** | 1.01 (0.59-1.72) |
| **Normal weight (18.5-25 kg/m^2^)** | 1.00 (reference) |
| **Overweight (25-30 kg/m^2^)** | 1.04 (0.60-1.79) |
| **Obese (>30 kg/m^2^)** | 1.18 (0.69-2.03) |
| **Missing** | 0.98 (0.56-1.72) |
| **Smoking Status** |  |
| **Never smoked** | 1.00 (reference) |
| **Ex-smoker** | 1.18 (0.96-1.45) |
| **Current smoker** | 1.21 (0.96-1.52) |
| **Data missing** | 1.20 (0.76-1.84) |
| **Exposure to selected infections*** | 1.50 (0.56-4.01) |
| **Exposure to selected medication**** | - |

SARS CoV-2=Severe Acute Respiratory Syndrome Coronavirus 2

* Epstein Barr virus (EBV), human cytomegalovirus (HCV), human herpesvirus 6 (HHV-6), human T lymphotropic virus type 1 (HTLV-1), hepatitis C virus, influenza A virus, and parvovirus B19

** procainamide, hydralazine, quinidine, and isoniazid
